# Community management: A model that effectively reduces rate of medication discontinuation in patients with schizophrenia

**DOI:** 10.1101/2025.04.23.25326268

**Authors:** Hua Ren, Qin Yang, Changjiu He, Jian Jiao, Zaiquan Dong

**Affiliations:** The Clinical Hospital of Chengdu Brain Science Institute, MOE Key Lab for Neuroinformation, School of Life Science and Technology, University of Electronic Science and Technology of China, Chengdu 610036, P.R. China; Sleep Medicine Center, Mental Health Center, Translational Neuroscience Center, West China Hospital, Sichuan University, Chengdu, P.R. China; Mental Health Center, West China Hospital, Sichuan University, Chengdu 610041, P.R. China

**Keywords:** community management, schizophrenia, discontinuation of treatment, adherence, compliance

## Abstract

**Introduction:** Patients with schizophrenia often experience high rates of medication interruptions, which have a range of serious consequences. Previous studies have shown that community management may help to address this situation.

**Objective:** To investigate treatment discontinuation in patients with schizophrenia under community management in China and analyze its associated factors.

**Methods:** The patients were assessed using a self-report questionnaire. The survey content included the following: treatment discontinuation, age, sex, education, residence, marital status, monthly family income, payment method for medication, employment, course of current disease, state of disease, insight, knowledge about drugs, types of drugs received, adverse effects of treatment, accepted follow-up, and family support.

**Results:** A total of 1531 patients with schizophrenia were included. Among them, 63 patients (4.1%) had discontinued medication at some point, while 1468 (95.9%) patients maintained treatment. The two groups showed statistically significant differences in payment methods for medication, state of disease, insight, knowledge about drugs, side effects of drugs, type of drugs received, and status of follow-up visits (*P* <0.05). The results of the multivariate logistic regression analysis showed that the period of disease onset, lack of insight, obvious side-effects of drugs, and lack of follow-up were risk factors for patients to discontinue medication (*P* <0.05).

**Conclusion:** Patients with schizophrenia under community management had a low rate of treatment discontinuation. The state of the disease, insight, side-effects of drugs, and status of follow-up visits are strongly related to medication interruption.

## 1 Introduction

Schizophrenia is a severe psychiatric disorder characterized by cognitive deficits, disordered thinking, emotional abnormalities, and multiple functional defects, with a high recurrence rate and significant harmful effects (1). Antipsychotics are essential for its treatment, but patient adherence is often low. Interrupting antipsychotic medications, even for a short time, reduces treatment effectiveness(2–4), increases the risk of relapse (5–8), and increases psychiatric hospital admissions(9, 10). Although continued medication is important, approximately 50% of patients with schizophrenia adhere to antipsychotic medications. Studies have suggested that effective medication management can reduce discontinuation risk(10), especially in community-based models that improve adherence and reduce violent behavior(11–14).

Community-based service models help patients maintain their connections with health care institutions and increase their access to community services (15). The main process of this working model is to ensure the continuity of patients’ treatment through continuous assessment and satisfaction of patients’ medical needs, so as to facilitate control of the disease, improve social function, reduce hospitalization and shorten hospitalization time, and improve the quality of life of caregivers and patients (16). Current models of community mental health services include community mental health care centers, day clinics, self-confident communities, and enhanced case management(17).

A case management model is a model commonly employed in community-based services with the ultimate goal of providing comprehensive mental health services to achieve individual goals and provide the most appropriate social functioning and a good quality of life in line with patients’ individual needs(18). This model has been shown to significantly promote clinical recovery, improve social and occupational functioning, and reduce length of hospital stay in patients with schizophrenia (15).

Since 2004, People’s Republic of China has implemented measures to manage severe mental disorders including schizophrenia (14). In 2009, the management of six major mental disorders, including schizophrenia, was integrated into the National Public Health service system (19). This is mainly based on the Assertive Community Treatment in America and Japan(20), with localized improvements. By 2025, >80% of counties are expected to offer extensive community rehabilitation services for mental disorders, with the aim of establishing a continuous prevention and treatment model primarily based on community management. Local health institutions provide services under this model, including health record management, follow-up, case management, medication guidance, and emergency medical assistance, with a focus on patients with schizophrenia. Although community-based services have been carried out in China for many years and have proved to be helpful for severe mental disorders, including schizophrenia(10), there is a lack of new evaluation on the implementation effect of community-based services in recent years, and especially the impact on reducing the medication discontinuation rate of patients has not been reported.

In the present study, we investigated the medication status of patients with schizophrenia under community management and analyzed related factors to verify that community-based management is an effective model for mental illness management that is well-suited to reduce medication interruptions, stabilize the condition, and reduce relapses, thereby improving the prognosis of individuals with schizophrenia.

## 2 Participants and Methods

### 2.1 Participants

The study surveyed registered patients with schizophrenia, managed by community administrators and meeting The International Statistical Classification of Diseases and Related Health Problems, 10^th^ revision (ICD-10) diagnostic criteria for schizophrenia from one urban district and one suburban district in Chengdu, China. This study was approved by the Ethics Committee on Biomedical Research in West China Hospital (approval number: 2020-797) and conducted in accordance with the Declaration of Helsinki. The survey was conducted from September 1 to 30, 2020, and 1,531 valid questionnaires were used. Prior to enrollment, formal verbal consent procedures were systematically implemented, with audio-recorded confirmation obtained from each participant. Patients were divided into the discontinuation and non-discontinuation groups based on whether medication was interrupted.

### 2.2 Methodology

Referring to previous studies(19, 21), the questionnaire was designed by experienced psychiatrists and included age, sex, education, residence, marital status, income, reimbursement ratio of drug insurance, employment status, disease course, disease state, insight, drug knowledge, drug type, side-effects, outpatient follow-up at the hospital, and family support.

- Medication discontinuation: The normative definition of medication discontinuation is difficult(22); in this study, it was defined as not taking any antipsychotic drugs for more than 15 days in the past month without a doctor’s advice or due to external factors like the coronavirus disease 2019 pandemic(23). Patients treated with long-acting injectable antipsychotics were excluded.
- Family income evaluation: The overall monthly income levels of major family members.
- Employment status: People who were able to work for most of the preceding year and had a fixed income were defined as employed; otherwise, they were considered unemployed.
- Disease state: This was divided into stable and unstable states, according to whether the disease state fluctuated or not.
- Insight: This was based on the cognitive understanding of the patients’ psychological state, and was divided into no, partial, or full insight.
- Knowledge about drugs: This was divided into three categories (good, average, and poor) according to the patient’s understanding of the importance of drug maintenance therapy, the effect of drug treatment, common side effects, and precautions for administration.
- Side-effects: This was based on the main complaints of patients and was divided into two categories (yes or no).
- Drug types: This referred to the kinds of different drugs taken by patients at the same time.
- Follow-up: Regular follow-up referred to patients who went to see the doctor according to medical arrangement or self-arrangement; intermittent follow-up referred to patients with irregular follow-up according to changes in their illness, although not according to the time agreed by their doctors; and no follow-up meant that there was neither a doctor’s appointment nor a self-arranged follow-up.
- Family support: This was based on family income that could or could not completely meet the patients’ medication expenses; we divided the types of family support into good, general, and poor.

### 2.3 Quality Control

The investigators were trained to ensure consistency and reliability in data collection and verification. The investigator fully explained the purpose of the study to the subjects in the presence of the main caregiver and completed the questionnaire individually. The questionnaire was collected onsite to ensure the authenticity and reliability of the data. The data were entered by one person and verified by a second person. Any inconsistencies in the data were verified by a third investigator.

### 2.4 Statistical Analysis

We initially described the differences between the medication discontinuation and non-discontinuation groups in terms of frequency (proportional composition) across general demographics and influencing factors. The statistical significance of the differences in these characteristics between the two groups was determined using the chi-squared test or Fisher’s exact test. We employed a binary logistic regression model to estimate the factors influencing medication discontinuation, with the significance level set at 0.05. The stepwise method was used to select covariates for inclusion in the multivariate logistic regression model. All statistical analyses were conducted using Statistical Analysis System (SAS) 9.4 software (SAS Institute Inc., Cary, North Carolina, America).

## 3 Results

### 3.1 Descriptive data of the included participants

A total of 1,531 patients with schizophrenia were enrolled in this study. The age at assessment ranged from 18 to 65 years for the medication discontinuation group (n=63) and from 18 to 77 years for the non-discontinuation group group (n=1468). The differences between the two groups were statistically significant, including payment methods for medication, state of disease, insight, knowledge about drugs, side effects, type of drugs taken (single or combined medication), and status of follow-up (all *P* <0.05; Table 1).

**Table 1.** Univariate analysis.

| Variable | Medication non-discontinuation group | Medication discontinuation group | $\chi^2$/degrees of freedom | *P* value |
| --- | --- | --- | --- | --- |
| Age (years), N (%) |  |  | 2.37/4 | 0.669 |
| ≤29 | 203 (96.2%) | 8 (3.8%) |  |  |
| 30–39 | 315 (96.0%) | 13 (4.0%) |  |  |
| 40–49 | 409 (96.5%) | 15 (3.5%) |  |  |
| 50–59 | 298 (96.1%) | 12 (3.9%) |  |  |
| ≥60 | 243 (94.2%) | 15 (5.8%) |  |  |
| Sex, N (%) |  |  | 0.22/1 | 0.641 |
| Female | 859 (96.1%) | 35 (3.9%) |  |  |
| Male | 609 (95.6%) | 28 (4.4%) |  |  |
| Education, N (%) |  |  | 0.87/2 | 0.647 |
| <Senior high school | 1086 (95.9%) | 46 (4.1%) |  |  |
| Senior high school | 222 (96.5%) | 8 (3.5%) |  |  |
| College or undergraduate | 160 (94.7%) | 9 (5.3%) |  |  |
| Residence, N (%) |  |  | 0.79/1 | 0.374 |
| Urban areas | 458 (95.2%) | 23 (4.8%) |  |  |
| Rural areas | 1010 (96.2%) | 40 (3.8%) |  |  |
| Marital status, N (%) |  |  | 4.26/2 | 0.119 |
| Married | 421 (94.8%) | 23 (5.2%) |  |  |
| Single | 830 (95.8%) | 36 (4.2%) |  |  |
| Divorced or widowed | 217 (98.2%) | 4 (1.8%) |  |  |
| Monthly household income, N (%) |  |  | 3.19/1 | 0.074 |
| <5000 | 1088 (95.4%) | 53 (4.6%) |  |  |
| ≥5000 | 380 (97.4%) | 10 (2.6%) |  |  |
| Payment, N (%) |  |  | 4.37/1 | 0.037 |
| Self-pay | 139 (92.7%) | 11 (7.3%) |  |  |
| Medical insurance | 1329 (96.2%) | 52 (3.8%) |  |  |
| Employment |  |  | 2.92/1 | 0.087 |
| No | 1122 (95.4%) | 54 (4.6%) |  |  |
| Yes | 346 (97.5%) | 9 (2.5%) |  |  |
| Disease course (year), N (%) |  |  | 1.62/2 | 0.446 |
| 0-5 | 390 (96.1%) | 16 (3.9%) |  |  |
| 5-10 | 466 (96.7%) | 16 (3.3%) |  |  |
| >10 | 612 (95.2%) | 31 (4.8%) |  |  |
| Disease state, N (%) |  |  | 31.54/2 | 0.000 |
| Unstable | 13 (86.7%) | 2 (13.3%) |  |  |
| Weak stable | 140 (88.1%) | 19 (11.9%) |  |  |
| Stable | 1315 (96.9%) | 42 (3.1%) |  |  |
| Insight level, N (%) |  |  | 51.67/2 | 0.000 |
| No | 19 (70.4%) | 8 (29.6%) |  |  |
| Partial | 510 (94.6%) | 29 (5.4%) |  |  |
| Full | 939 (97.3%) | 26 (2.7%) |  |  |
| Drug knowledge, N (%) |  |  | 21.42/2 | 0.000 |
| Poor | 142 (89.3%) | 17 (10.7%) |  |  |
| Generally | 894 (96.1%) | 36 (3.9%) |  |  |
| Good | 432 (97.7%) | 10 (2.3%) |  |  |
| Side effects, N (%) |  |  | 21.16/1 | 0.000 |
| No | 1249 (96.9%) | 40 (3.1%) |  |  |
| Yes | 219 (90.5%) | 23 (9.5%) |  |  |
| Medication type, N (%) |  |  | 7.17/1 | 0.007 |
| Single | 402 (93.7%) | 27 (6.3%) |  |  |
| Combined | 1066 (96.7%) | 36 (3.3%) |  |  |
| Follow-up, N(%) |  |  | 43.42/2 | 0.000 |
| No | 221 (88.4%) | 29 (11.6%) |  |  |
| Intermittent | 637 (96.8%) | 21 (3.2%) |  |  |
| Regular | 610 (97.9%) | 13 (2.1%) |  |  |
| Family support, N (%) |  |  | 5.15/2 | 0.076 |
| Poor | 75 (93.8%) | 5 (6.3%) |  |  |
| General | 532 (94.7%) | 30 (5.3%) |  |  |
| Good | 861 (96.9%) | 28 (3.1%) |  |  |

### 3.2 Factors associated with medication discontinuation

Univariate and multivariate regression analyses were done with the interruption of medication as the dependent variable (discontinuation = 0; non-discontinuation = 1), and age, sex, education, residence, marital status, income, payment methods, employment, disease course, disease state, insight, drug knowledge, medication types, side effects, follow-up, and family support as the independent variables. The results of multivariate logistic regression analysis showed that the period of disease onset, lack of insight, obvious side effects of drugs, and lack of follow-up were risk factors for patients to discontinue medication (all *P* <0.05; Table 2).

**Table 2.**
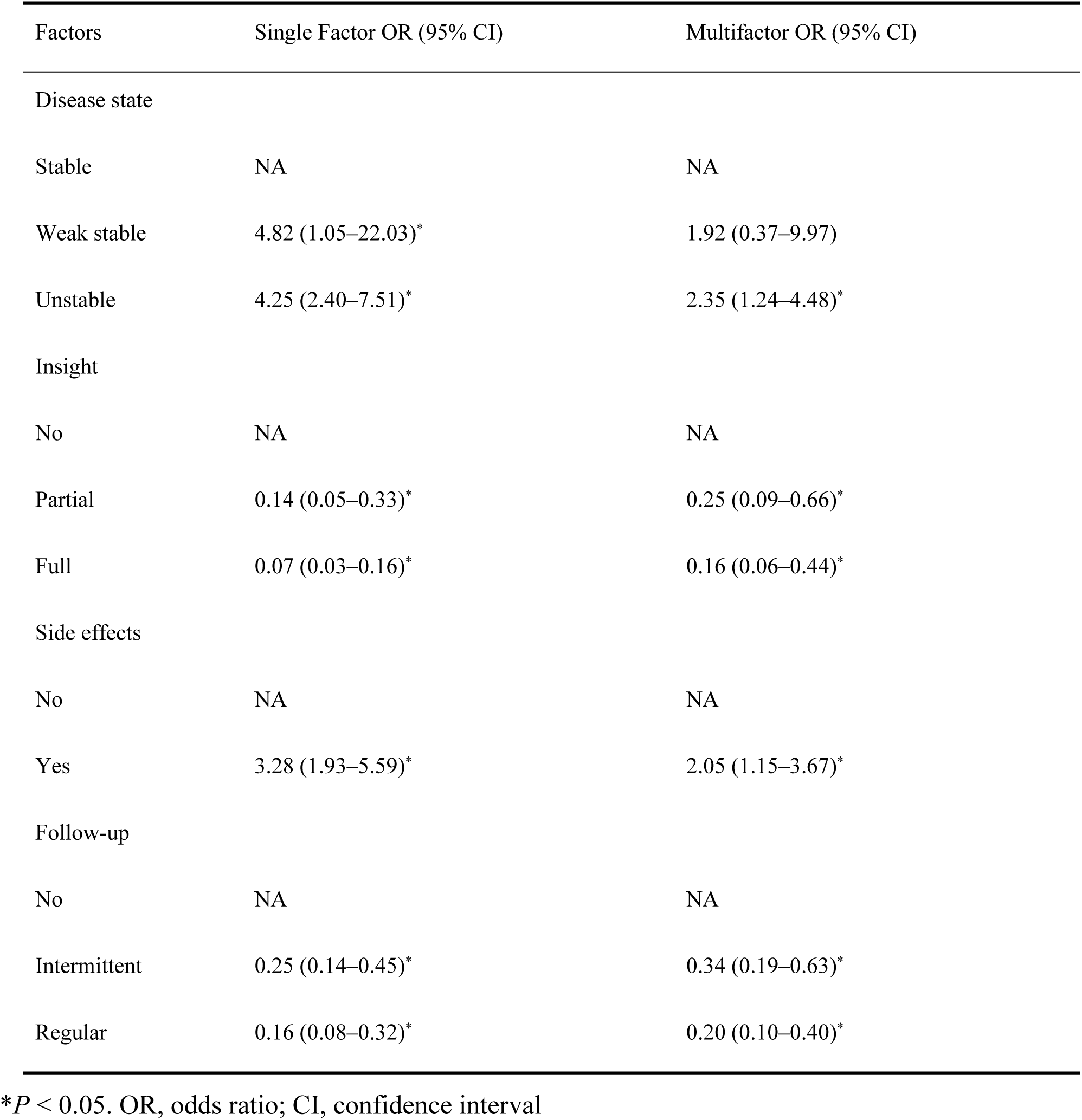
Results of the multivariae meta-regression analysis.

| Factors | Single Factor OR (95% CI) | Multifactor OR (95% CI) |
| --- | --- | --- |
| Disease state |  |  |
| Stable | NA | NA |
| Weak stable | 4.82 (1.05–22.03)* | 1.92 (0.37–9.97) |
| Unstable | 4.25 (2.40–7.51)* | 2.35 (1.24–4.48)* |
| Insight |  |  |
| No | NA | NA |
| Partial | 0.14 (0.05–0.33)* | 0.25 (0.09–0.66)* |
| Full | 0.07 (0.03–0.16)* | 0.16 (0.06–0.44)* |
| Side effects |  |  |
| No | NA | NA |
| Yes | 3.28 (1.93–5.59)* | 2.05 (1.15–3.67)* |
| Follow-up |  |  |
| No | NA | NA |
| Intermittent | 0.25 (0.14–0.45)* | 0.34 (0.19–0.63)* |
| Regular | 0.16 (0.08–0.32)* | 0.20 (0.10–0.40)* |

\**P* < 0.05. OR, odds ratio; CI, confidence interval

## 4 Discussion

The need for supervised management of drug use in patients with schizophrenia is obvious. Leaving medication solely to patient discretion or guardian supervision would be disadvantageous, given the mixed public perception of the disease. A community-based management model may be an effective method to reduce the probability of medication interruption in patients with schizophrenia.

The present study, conducted in Chengdu, showed that the medication discontinuation rate in patients with schizophrenia under community management was 4.1%, which was lower than that in general patients with schizophrenia (9, 24–26). A study with 4 years of follow-up in the United States of America showed that 36% of patients had poor adherence to medication each year and 61% of patients discontinued their medication at some point during the follow-up period (27)(22). Combined with current reports, the average adherence rate of patients with schizophrenia is approximately 40%–60%, and some studies have reported that 75%–90% of patients become nonadherent within 1– 2 years after discharge. Valenstein et al. reported that effective management measures can significantly reduce the probability of medication interruption (28).This suggests that the factors at play may include supervision of medication, facilitation of access to medication, more opportunities for follow-up visits, and humanistic care from community managers. The low rates of medication discontinuation in the present study may be due to the stricter standard for the definition of medication discontinuation (interruption of antipsychotic drugs for 30 d). In some studies, medication discontinuation was defined as the administration of an antipsychotic drug for 14 d(29).

The results of the multivariate logistic regression analysis showed that the discontinuation rates of medication in the period of disease onset were 2.35 times lower than those in the stable disease stage (95% confidence interval: 1.24–4.48). It may be due to the fact that patients in the acute phase are controlled by psychotic symptoms such as hallucinations and delusions, are unable to properly assess their mental state, and refuse to take medication(29–31). Thus, a stable disease state is necessary to reduce medication interruptions. One study showed that patients with schizophrenia had high medication adherence when their disease was in the normal-to-mild stages (*P* = 0.015)(32). The severity of symptoms and the ability to recognize psychotic symptoms are independent risk factors for medication adherence(33).

Second, the lack of disease insight was a risk factor for medication discontinuation, which is consistent with the conclusions of most studies(34–36). Good insight into the disease implies a proper understanding of the disease state and its adverse consequences, which is a crucial marker of disease recovery and an important condition for medication adherence. Consequently, improving patient’s insight into the disease is a major goal in schizophrenia treatment.

Furthermore, the obvious side-effects of drugs are also an important cause of medication interruption, as indicated in previous studies(7, 24, 34, 37). This study did not investigate the types of medications taken by the patients, such as first generation and second generation antipsychotics. Research has shown that second generation antipsychotics which often have milder side effects may enhance medication compliance and reduce discontinuation(7, 38).

Lastly, regular follow-up was also one of the factors related to medication discontinuation. While there is limited research on this aspect, it could be beneficial to adjust medications in a timely manner through regular follow-up. Information about the disease, health education, and emotional support provided during follow-up visits may help patients to better adhere to their medication regimens.

In summary, despite challenges such as a lack of professional talent and low cooperation from patients or their families in community mental health management, the rate of medication discontinuation among patients with schizophrenia under community management is significantly lower than that in the general population. The risk factors leading to discontinuation are unstable disease status, lack of insight, severe side effects of drugs, and lack of regular follow-up, highlighting the key areas of community management for patients with schizophrenia. Additionally, studies have indicated that poor family relationships and support systems are significant causes of medication discontinuation(7), and robust social support could help reduce relapse in patients with schizophrenia(39). However, the present study did not reach the same conclusion, suggesting that community management may play a partial role as a support system for patients. Moreover, cognitive-behavioral therapy and other socio-psychological interventions can reduce the risk of rehospitalization and are effective in enhancing adherence and reducing medication discontinuation (7).

This study had several limitations. First, it did not consider differences in the types of medication, as studies have shown variations in discontinuation rates between first- and second-generation antipsychotics (40). Second, it did not account for the effectiveness of the medication, particularly the subjective effects experienced by patients, which requires further clarification in future research. Third, objective assessments of medication discontinuation, such as blood or urine medication levels, were not performed, which may have caused bias.

## Data Availability Statement

The data is available upon reasonable request by the corresponding author.

## Author Contributions

ZD and YY received the funding and designed the study. HR analyzed the data together with YQ, JJ and CH. HR drafted the manuscript. All authors were involved in this study, and have read and approved the final version of the manuscript.

## Acknowledgments

We would like to thank the participants of this study and the medical staff who supported the data collection. We would also like to thank Editage for English language editing.

